# Long-read Oxford Nanopore sequencing reveals complex rearrangements and regulatory disruption in malignant pleural mesothelioma

**DOI:** 10.64898/2026.05.26.26353864

**Authors:** Mazin H. Alhazmi, Charlotte Poile, Joanna Dzialo, Aleksandra Bzura, Kudzayi Kutywayo, Dean Fennell, Edward J. Hollox

## Abstract

Malignant pleural mesothelioma (MPM) is a rare malignancy characterised by extensive structural genomic alterations and a low burden of recurrent single nucleotide variants. However, the full spectrum and functional impact of structural variation (SVs) remain incompletely understood because short-read sequencing has limitation ability to resolve complex genomic rearrangements. Here, we performed integrated short-read and long-read whole-genome sequencing on tumour-normal pairs from three MPM patients, together with RNA sequencing and nanopore-derived promoter methylation profiling. Long-reads sequencing substantially improved SV detection, identifying 61-156 novel SVs per sample, including complex rearrangements and breakpoint-resolved events affecting cancer-associated genes. Complex SV clusters consistent with chromoplexy and chromothripsis were observed and frequently involved oncogenes. Integration with transcriptomics data showed that several SVs-affected genes, including *WEE1* and *GPC6*, exhibited increased expression independent of gene dosage. Promoter methylation analysis revealed a conserved bimodal methylation landscape across tumours and a significant inverse relationship with gene expression. SV-associated genes showed coordinated promoter hypermethylation and transcriptional activation, suggesting that SVs may influence gene regulation through epigenetic mechanisms. Survival analysis using the TCGA-MESO cohort further showed that elevated expression of *WEE1* and *GPC6* was associated with poorer overall survival. Together, these findings highlight the value of long-read sequencing for uncovering functionally and clinically relevant structural variation in MPM.

## Introduction

Malignant pleural mesothelioma (MPM) is a rare malignancy arising from the mesothelial lining of the pleura, primarily linked to asbestos exposure [1,2]. Despite advances in diagnostic imaging and multimodal treatment strategies, individuals with MPM often exhibit poor clinical outcomes, with a median overall survival of less than one year [3]. At the molecular level, MPM is characterised by a low overall single nucleotide mutational burden and a paucity of recurrent single mutations, distinguishing it from many other solid tumours [4]. Instead, structural variants (SVs), including somatic copy number alterations (SCNAs), dominate the genomic landscape of MPM [4,5,6]. However, the full extent and impact of these rearrangements remain poorly understood, in large part due to limitations of traditional short-read sequencing in reliably assaying the full spectrum of SVs [7].

Short-read whole genome sequencing (WGS) has enabled genome-wide identification of somatic alterations in MPM, including frequent inactivation of tumour suppressor genes such as *BAP1, CDKN2A, NF2* and *LATS2* [4,5,6,8,9,10]. Nevertheless, short-read approaches are inherently limited in resolving large or complex SVs, particularly those involving repetitive or low complexity regions, or events such as large insertion, inversion, and translocation with imprecise breakpoint [7,11]. These challenges lead to high false negative rates of SV detection and imprecise SV breakpoint resolution, hampering efforts to link SVs to gene dysregulation or clinical phenotypes. As a result, many potentially pathogenic rearrangements in MPM may remain undetected, contributing to gaps in our understanding of tumorigenesis and disease progression [12].

The emergence of long-read sequencing technologies, such as Oxford Nanopore Technologies (ONT), offers a powerful alternative for resolving complex genomic rearrangements [13]. Long-read WGS allows for direct observation of structural events spanning tens to hundreds of kilobases, enables more accurate SV detection and phasing of allelic variants [14,15,16]. While long-read sequencing has been applied in select cancer types [17], its use in MPM remains limited, and comparative benchmarking of SV detection tools in this context is lacking. Moreover, few studies have integrated long-read based SV discovering with multi-omics data such as gene expression, methylation and clinical outcomes to assess the functional relevance of novel SVs [17].

SVs have the potential to impact gene regulation not only by direct disruption of coding regions but also by alteration of cis-regulatory elements, topologically associated domains, and promoter methylation states [18]. Therefore, some SVs may modulate gene expression beyond direct copy number alteration of genes, yet their transcriptional consequences in MPM are largely unexplored [4,17]. Furthermore, the clinical relevance of SV-affected genes including their prognostic value remain unclear, despite increasing interest in identifying novel biomarkers and therapeutic targets in MPM [19].

In this study, we applied whole genome long-read and short-read genome sequencing to matched tumour-normal pairs of three MPM patients to comprehensively profile somatic SVs. We compared the performance of two long-read SV callers against SVs called by the short-read SV caller Manta SVs, and with RNA expression, copy number, and promoter methylation data to identify candidate genes whose regulation may be altered by complex rearrangements. Expression-based survival analysis in the TCGA-MESO cohort further assessed the clinical relevance of SV-affected genes. Finally, selected SV events were validated experimentally by PCR amplification and Sanger sequencing.

Together, our findings show the importance of long-read sequencing in fully revealing novel SVs in MPM, aiding in understanding the full mutational landscape of the MPM genome. These novel SVs include complex large-scale chromoplexy events as well as smaller insertions and deletions, and can affect known driver genes. Long-read genomic sequencing also reveals genomewide methylation patterns, allowing deconvolution of cell type mixtures and analysis of differential methylation at key cancer genes.

## Material and methods

### Patient sampling

This research was approved by NHS research ethics committees references 14/LO/1527 and 14/EM/1159. Patients with a confirmed histological diagnosis of MPM were recruited into the MEDUSA cohort, scheduled for extended pleurectomy decortication at Glenfield Hospital (University of Leicester). Tumour samples (MED239, MED241 and MED243) were collected from the pleura, with no specific region at surgery, and peripheral blood was collected at the same time.

### Nucleic acid extraction

Low molecular weight DNA from tumour tissue was semi-automatically extracted using the Maxwell® RSC and Authentication Kit (Promega Corporation, Madison, WI) according to the manufacturer’s protocol. Approximately 20⍰mg of tumour tissue was used for extraction. Peripheral blood was collected in 5⍰mL EDTA tubes prior to each end-pleural drainage (EPD) procedure. Non-tumour DNA was extracted from 200⍰μL of whole blood using the Maxwell® RSC and Authentication Kit (Promega Corporation, Madison, WI), according to the manufacturer’s instructions. High-molecular-weight (HMW) DNA was extracted using the Monarch® HMW DNA Extraction Kit for Tissue (New England Biolabs, UK), according to the manufacturer’s protocol. Approximately 10– 25⍰mg of tissue was used per extraction. DNA concentration was measured using the Qubit™ High Sensitivity (HS) DNA concentration and purity were assessed using the Qubit™ HS DNA Assay Kit (Invitrogen) and NanoDrop™ 2000 spectrophotometer (Thermo Scientific), respectively. After processing, low-molecular weight DNA samples were stored at -20 °C, and HMW DNA samples stored at 4 °C.

Total RNA was extracted from MPM tumour tissues using the RNeasy Mini Kit (Qiagen, Germany), following the manufacturer’s instructions. Approximately 50⍰mg of tissue was used per extraction. RNA concentration, integrity (RNA Integrity Number; RIN), and purity (A260/A280 ratio) were assessed using the Agilent 2100 Bioanalyzer with the RNA 6000 Nano Chip. All samples had a RIN⍰> ⍰8.

### Short-read whole genome sequencing and analysis

Genomic DNA from mesothelioma tissues and matched blood samples was sequenced using 150⍰bp paired-end sequencing on the Illumina NovaSeq 6000 (MED239) and NovaSeq X Plus (MED241 and MED243) platforms at 50x coverage, and the matched non-tumour samples were sequenced at 30x coverage. All sequencing was conducted by Novogene Ltd (Cambridge, UK). The quality of the FASTQ paired-end WGS and WES files was assessed using parameters such as per-base sequence quality, per-sequence quality scores, over-represented sequences, and adapter content using FastQC v0.11.5 [20]. Then, quality trimming, trimmomatic was used [21]. The paired-end reads were aligned with the human genome reference (hg38) available from the 1000 Genomes Project (GCA_000001405.29) using BWA v0.7.17 [22]. The alignment files were converted into BAM files and sorted and indexed using SAMtools v1.9 [23]. Artefactual duplicate reads were removed using Picard v2.6.0 [24]. Tumour purity was estimated using Theta2 [25]. Somatic SNVs were identified using Mutect2 v4.1.0.0 [26]. CNVkit v0.9.13 [27] was used to detect somatic CNVs from WGS tumour-normal paired samples. Manta v1.4.0 [28] was used to identify the somatic SVs in MPM tumour-normal samples.

### Long-read whole genome sequencing and analysis

5 μg of HMW DNA of MPM tissues and blood were used to generate long-read sequencing ≥ 20X. The sequencing was performed using PromethION platform, with library preparation done using Nanopore genomic DNA ligation sequencing SQK-LSK114 chemistry (ONT®) and PromethION Flow Cell R10.4.1 flow cells (ONT®). All sequencing was conducted by Novogene Ltd (Cambridge, UK).

Raw signal data in FAST5 format were converted to nucleotide sequence in FASTQ format using Guppy v6.1.2. Basecalling was performed using the configuration file dna_r10.4.1_450bps_hac_prom.cfg, with ha_5mc used for methylation calling based on the Flow Cell (R10.4.1) and Ligation Sequencing Kit used (SQK-LSK114) to generate the FASTQ files. NanoPlot (v1.38.1) was used to check the quality of the sequencing reads and sequencing alignment [29]. Long-read were aligned to the human genome reference (GRCh38.p14 Primary Assembly) available from the 1000 Genomes Project (GCA_000001405.29) using Minimap2 (v2.24) [18]. The alignment files were converted into BAM files and sorted and indexed using SAMtools v1.9 [23].

### SNVs calling

NanoCaller [30] is a computational tool designed to detect small variants, including single-nucleotide variants (SNVs) and small insertions and deletions (indels), from long-read sequencing data. In this study, long-read whole-genome sequencing (WGS) data from malignant pleural mesothelioma (MPM) tumour tissue and matched normal (blood) samples were analysed using NanoCaller to identify SNVs. BAM files for tumor and matched normal samples were processed independently using the human reference genome (hg38; 1000 Genomes Project, GCA_000001405.29). This step generated variant calls in variant call format (VCF).

To complement this approach, variant calling was also performed using ClairS (v0.4.5) [31], a deep learning-based method optimised for somatic SNV detection from long-read sequencing data. ClairS incorporates tumour–normal paired analysis to distinguish somatic variants from germline variation, providing an alternative framework for SNV identification.

For NanoCaller-derived variants, somatic SNVs were inferred by comparing tumour and matched normal VCF files using bcftools [23], retaining variants present exclusively in tumour samples. Variant calls from both NanoCaller and ClairS were subsequently filtered to retain high-confidence SNVs. To prioritise functionally relevant mutations, variants were restricted to those located within exonic regions by intersecting variant coordinates with reference gene annotations.

### SVs calling

Sniffles [15] and Severus [16] were used to detect SVs from MPM tumour match normal (blood) sample using the following parameters: threads 10, minimum map quality 20, minimum SV length 50 bp, and minimum read support 3. SVs were detected using Sniffles in tumour and matched normal samples separately. Then Survivor v1.0.7 [32] was used to distinguish the somatic SVs from germline. Severus is an alternative SV detection tool designed to identify somatic SVs using a phased breakpoint graph approach. Two main files exist that need to be prepared to detect SVs using Severus: (1) phased SNPs from matched normal sample and (2) the haplotagged.bam file for both normal and tumour. Here, NanoCaller was used to identify and phase the SNPs from the BAM files for the normal sample, and WhatsHap v2.8 [33] was used to add haplotype tags to the BAM files. Finally, AnnotSV v3.1.3 [34] was used to annotate the SV VCF files.

For benchmarking of tools, Minda v0.0.2 [16] tools were used to assess the performance of Severus and Sniffles for somatic SV detection using long-read WGS, comparing their results against a truth set generated from Manta somatic SV calls from the short-read WGS data.

For validation of SVs, PCR primers were designed to span breakpoint junctions and were validated by PCR using standard conditions and Sanger Sequencing (Source Biosciences, Nottingham) on tumour genomic DNA, using genomic DNA from matched blood as a negative control (S1 Table).

### Methylation analysis

Base-modified reads were processed using modbam2bed v0.10.0 [35] with --5mc flag and –CpG flags to extract CpG methylation calls. differential methylation analysis was performed using methylKit v0.6 [36] on MPM tumour samples. Promoter-level methylation differences between positive SVs and negative SVs were assessed.

### RNA sequencing and transcriptional count

Total RNA libraries of MPM tumour samples were prepared and sequenced by Novagene using Illumina NovaSeq6000 and NovaSeq X Plus platforms. Raw sequence data (in FASTQ format) was quality checked using FastQC v0.11.5. Transcript abundance was quantified using Salmon [37] in quasi-mapping mode. Expression values were reported in transcripts per million (TPM).

### Assessment of the impact of the novel structural variants

To investigate the potential clinical relevance of the transcriptional changes associated with novel SVs, a three-step validation workflow was implemented using the TCGA Malignant Pleural Mesothelioma (TCGA-MESO) cohort via the BioLinkTCGA R package (v3.21; v3.4.1) [38]. In the first step, three tumour samples from this study were identified as harbouring novel SVs that disrupted specific genes. RNA sequencing data were used to compare the expression of each SV-affected gene between SV-positive and SV-negative samples. This analysis yielded a shortlist of candidate genes with expression potentially influenced by the presence of SVs. In the second step, these candidate genes were queried in the TCGA MPM RNA sequencing dataset via BioLinkTCGA to assess their expression profiles in a larger, clinically annotated cohort. For each gene, the median TPM value across all TCGA samples was computed and used to categorise patients into high- and low-expression groups.

In the final step, Kaplan–Meier survival analysis was performed to determine whether gene expression levels were associated with overall survival. Log-rank tests were applied to assess statistical significance, and the Benjamini–Hochberg (BH) method was used to correct for multiple testing. Genes with an adjusted p-value < 0.05 were considered statistically significant.

## Results

### Sequencing Data Summary

We generated whole-genome long-read, short-read sequence and total RNA sequencing data from three MPM patients. MED239 and MED241 were isolated from the right side, while MED243 was isolated from left side (S2 Table). We obtained over 130 billion bases (>20X in depth) long-read data per sample with a mean read N50 of 24 kb (range from 20 kb to 28 kb). The maximum read length was between 580 kb and 248 kb across all samples. With Minimap2, > 99% of the reads were mapped to the reference genome (GRCh38) (Table 1, S3A-D Tables). These metrics demonstrate the suitability of long-read WGS for resolving structural variation in MPM genomes. Short-read WGS achieved a mean coverage of the mapped reads for tumour samples ranging from 47.7X to 43.9 X (Table 1). In addition, RNA sequencing generated over 12 Gb of data per sample, providing sufficient depth for transcriptomic analyses (Table 1).

**Table 1:** Summary of the WGS long-read, WGS short-read and RNA sequencing of MPM patient samples.

|  |  | **MED239** |  | **MED241** |  | **MED243** |  |
| --- | --- | --- | --- | --- | --- | --- | --- |
|  |  | **Tumour** | **Blood** | **Tumour** | **Blood** | **Tumour** | **Blood** |
| Long-read<br>WGS | **N50 (kb)** | 20 | 18 | 24 | 21 | 28 | 25 |
|  | **Total bases (Gb)** | 119.1 | 144.3 | 130.8 | 120.6 | 122 | 131.6 |
|  | **Estimated coverage (fold)** | 38 | 46 | 41 | 38 | 39 | 42 |
|  | **Longest read length (kb)** | 580 | 601 | 482 | 256 | 248 | 687 |
|  | **Mapping percentage** | 99.78 | 99.82 | 99.42 | 99.43 | 99.67 | 99.67 |
| Short-read<br>WGS | **sequencing coverage (fold)** | 48.2 | 40 | 47.9 | 29.3 | 46.3 | 28.3 |
|  | **Mean coverage of mapped read (fold)** | 47.7 | 39.5 | 47.2 | 28.9 | 43.9 | 27.9 |
|  | **Percentage of reads mapped** | 99.91 | 99.91 | 99.93 | 99.94 | 99.21 | 99.95 |
| RNA<br>Sequencing | **Total bases (Gb)** | >12 | N/A | >12 | N/A | >12 | N/A |

### Somatic single nucleotide variation analysis

To evaluate concordance of somatic single nucleotide variation (SNV) detection between sequencing platforms, variant calling was performed using both short-read and long-read WGS data. Using Mutect2, we identified approximately 2,854–4,208 somatic single nucleotide variants (SNVs) per tumour from short-read whole-genome sequencing (WGS) (Table 2). Of these, 1,041–1,802 were missense mutations, while 132–560 were stop-gained variants, indicating potential functional relevance. A total of 23 missense SNVs at identical genomic coordinates were shared between a minimum of two samples, and, of these, seven missense SNVs overlapped with those reported in the COSMIC cancer mutation database, implicating known MPM-associated loci (S4 Table). In contrast, no stop-gained SNVs were recurrent at the same genomic position across samples. At the gene level, 353 genes harboured missense SNVs in two or more samples, 127 of which overlapped COSMIC. Additionally, three genes with recurrent stop-gained variants were identified across patients, although none overlapped COSMIC (S4 Table). Overall, these results indicate a relatively low SNV burden and limited recurrence, consistent with a genomic landscape predominantly driven by structural and copy number alterations rather than point mutations.

**Table 2.** Comparison of Somatic SNVs calls from short-reads WGS and long-read WGS across MPM tumour samples.

| Sample | Caller | Total SNVs | Missense | Stop-gained | Total SNVs overlapped with Mutect2 | Missense overlapped with Mutect2 | Stop-gained overlapped with Mutect2 |
| --- | --- | --- | --- | --- | --- | --- | --- |
| **MED239** | Mutect2 | 2,854 | 1,041 | 56 | — | — | — |
|  | NanoCaller | 8,523 | 516 | 75 | 100 | 3 | 0 |
|  | ClairS | 263 | 79 | 4 | 35 | 17 | 1 |
| **MED241** | Mutect2 | 3,483 | 1,266 | 59 | — | — | — |
|  | NanoCaller | 6,394 | 443 | 60 | 84 | 0 | 0 |
|  | ClairS | 197 | 57 | 12 | 52 | 28 | 3 |
| **MED243** | Mutect2 | 4,208 | 1,802 | 132 | — | — | — |
|  | NanoCaller | 2,577 | 299 | 12 | 26 | 1 | 0 |
|  | ClairS | 300 | 76 | 2 | 36 | 15 | 0 |

In contrast, long-read using variant calling NanoCaller and ClairS identified a broader range of candidate somatic SNVs across all samples (Table 2). Nanocaller identified 2,577-8,523 SNVs, including 299-516 missense variants and 12-75 stop-gained variants, whereas ClairS identified substantially fewer variants (197-300 somatic SNV per sample), reflecting differences in caller sensitivity and specificity.

Although long-read WGS identified larger number of candidates SNVs across all samples compared to short-read WGS, only small subset of variants (26-100 per sample) was consistently detected by both methods (Table 2). These finding indicate that, in this dataset, long-read sequencing yields a larger set of candidates SNVs but with limited overlap with short-read calls. Gavin that short-read WGS using Mutect2 represents the current clinical standard for somatic SNVs detection, and only SNVs identified using Mutect2 were used in subsequent analysis.

### Copy Number Alteration Analysis

Somatic copy number alterations (SCNAs) were identified using CNVkit applied to short-read WGS data. Across the three MPM samples, between 485 and 625 SCNA events were detected, with an average event size ranging from 813.9⍰kb to 1.1⍰Mb (S5 Table). Tumour purity, estimated using Theta2, was between 0.49 and 0.50 across all samples (S6A-C Figures).

The GISTIC2 algorithm was applied to identify statistically significant recurrent SCNAs at the chromosome arm level across the MPM cohort. WGS data from three tumour samples, each with matched normal controls, were analysed to distinguish somatic events and assess recurrent patterns of genomic alteration. The results are summarised in a heatmap representation (Figure 1), illustrating the distribution of arm-level SCNAs across samples.

**Figure 1.**
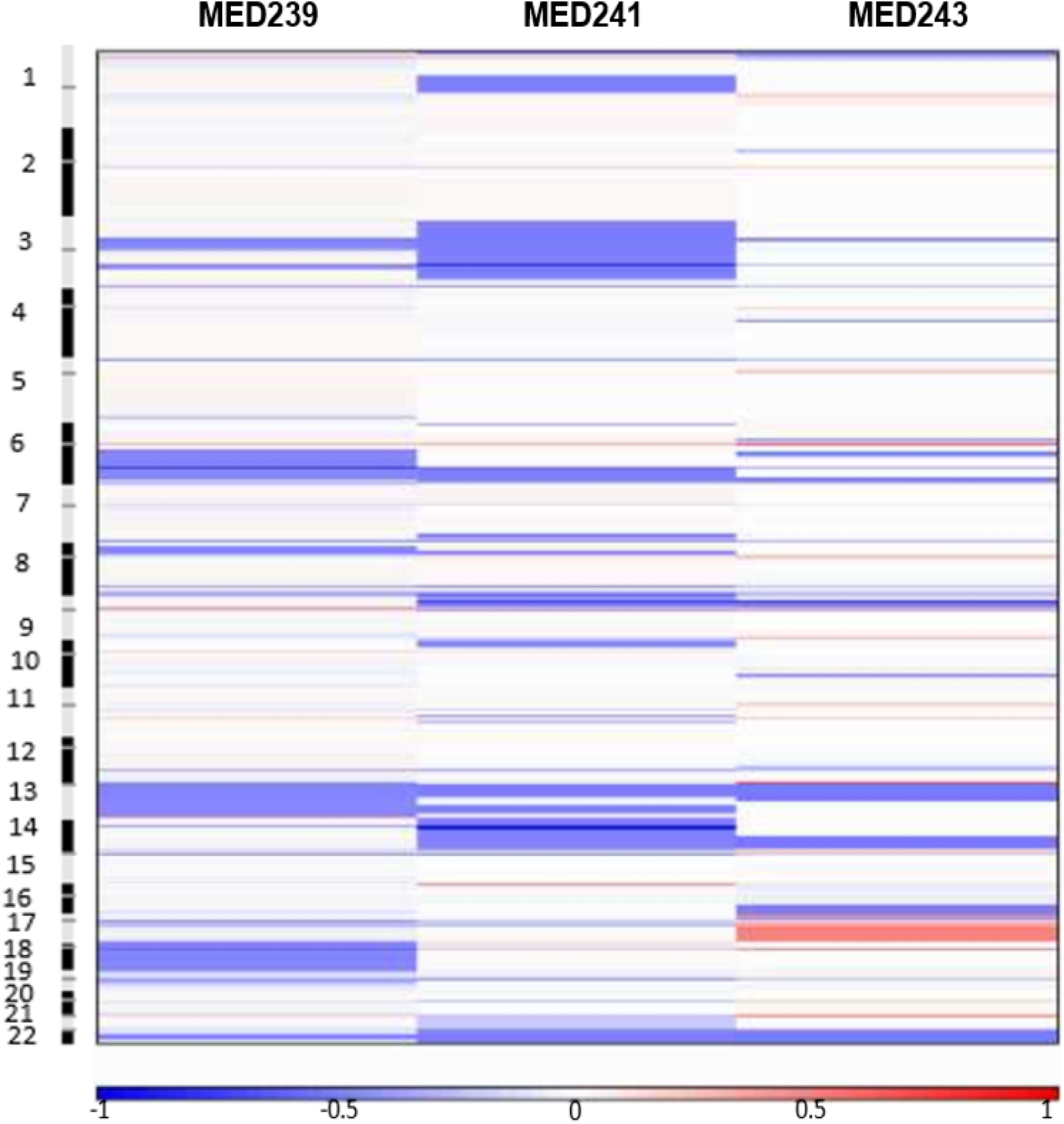
Frequency of recurrent arm level CNAs: The chromosomes are displayed along the y-axis, with each row representing a different chromosome arm, while the x-axis corresponds to specific genomic loci in specific samples. The colour intensity in these regions is proportional to the magnitude of the copy number changes, with deeper reds indicating more arm-level amplification and deeper blues indicating more arm-level deletion

Recurrent alterations were predominantly characterised by copy number losses rather than gains. In MED239, frequent losses were observed on chromosome arms 13q, 6q, and 18. MED241 exhibited losses across multiple regions, including chromosomes 3, 14, 9p, and 22. In contrast, MED243 showed a more mixed profile, with recurrent losses at 22q alongside gains at 13p and 17q (Figure 1). Overall, these findings highlight a predominance of arm-level deletions across MPM tumours, consistent with widespread loss of genomic material as a defining feature of the disease.

### Comparison of SVs detection between short-read and long read WGS

We identified SVs affecting MPM driver genes using long read sequencing ONT sequencing using Sniffles and Severus. For the high-sequence coverage MPM patient dataset, with cutoffs of a minimum SV length of 50 bp and a minimum of two supporting reads, Sniffles and Severus identified approximately 151 to 1,688 balanced and unbalanced somatic SVs (S7 Table). Sniffles called the highest number of somatic SVs among the MPM patient samples, ranging from 176 to 1,688, while Severus called the lowest number of somatic SVs, ranging from 151 to 243 (S8.A-B Table).

Five SV-types were identified in each sample analysed by Sniffles and Severus: insertion (INS), deletion (DEL), Duplication (DUP), inversion (INV), and translocation (BND). The most common SV types detected by Sniffles were Insertion and DEL (∼54.3% and ∼41.6%, respectively), followed by BND at ∼1.52%. The lowest SVs were DUP and INV (∼0.5% and ∼1%, respectively). In contrast, Severus identified DEL and BND as the most common SV types (∼42.6% and ∼26%, respectively), followed by INV at ∼16.5%, while INS and DUP were least frequently observed SV types (∼11.3% and ∼3.4%, respectively) (S8.A-B Table and S8.C Figure).

To evaluate the accuracy and concordance of long-read SV calls, we benchmarked Sniffles and Severus against Manta using the Minda evaluation tool, calculating true positives (TP), false positives (FP), false negatives (FN), precision, recall, and F1 scores (Table 3). Across all three samples, Severus consistently outperformed Sniffles in terms of precision and F1-score. Notably, Sniffles exhibited high sensitivity but lower precision, likely due to overcalling in repetitive regions or less stringent filtering of low-confidence events. In contrast, Severus provided a more conservative but accurate SV callset, achieving higher F1-scores across samples.

**Table 3:**
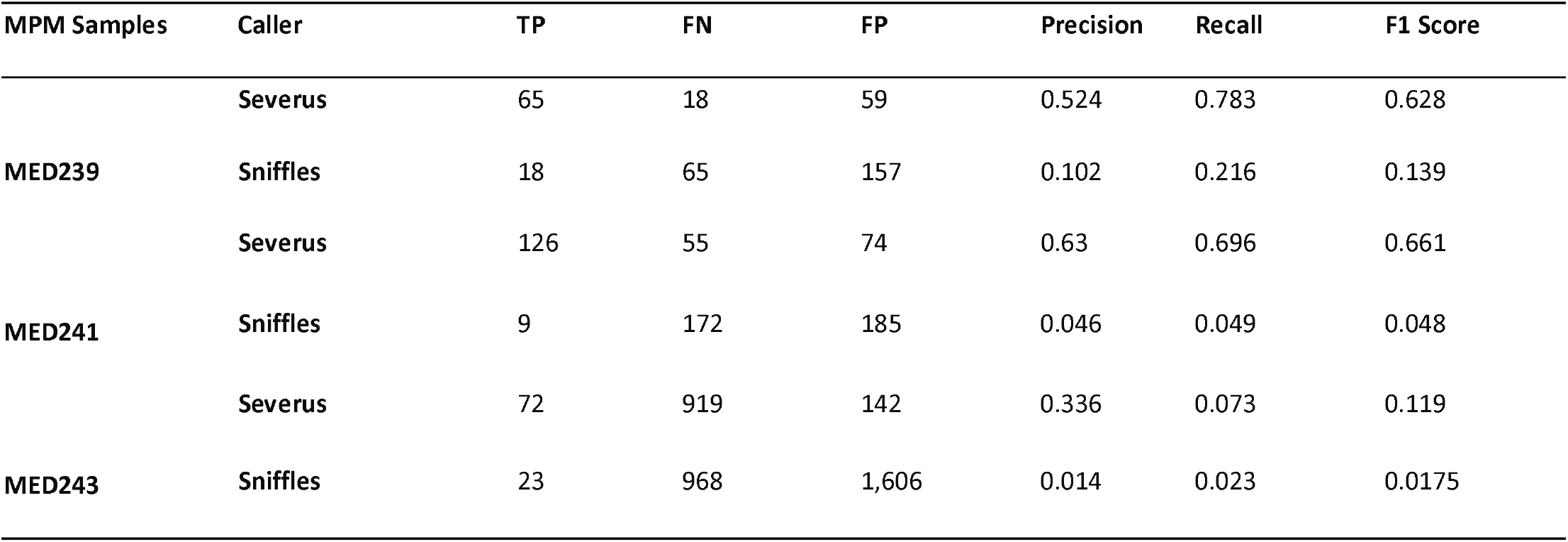
Long-read SV Caller Performance.

Importantly, Severus identified between 61 and 156 novel SVs not detected by Manta (Table 4). These long-read-exclusive SVs were enriched for deletions and insertions, underscoring the limitations of short-read sequencing in capturing complex or repetitive structural rearrangements (S9 Table).

**Table 4:**
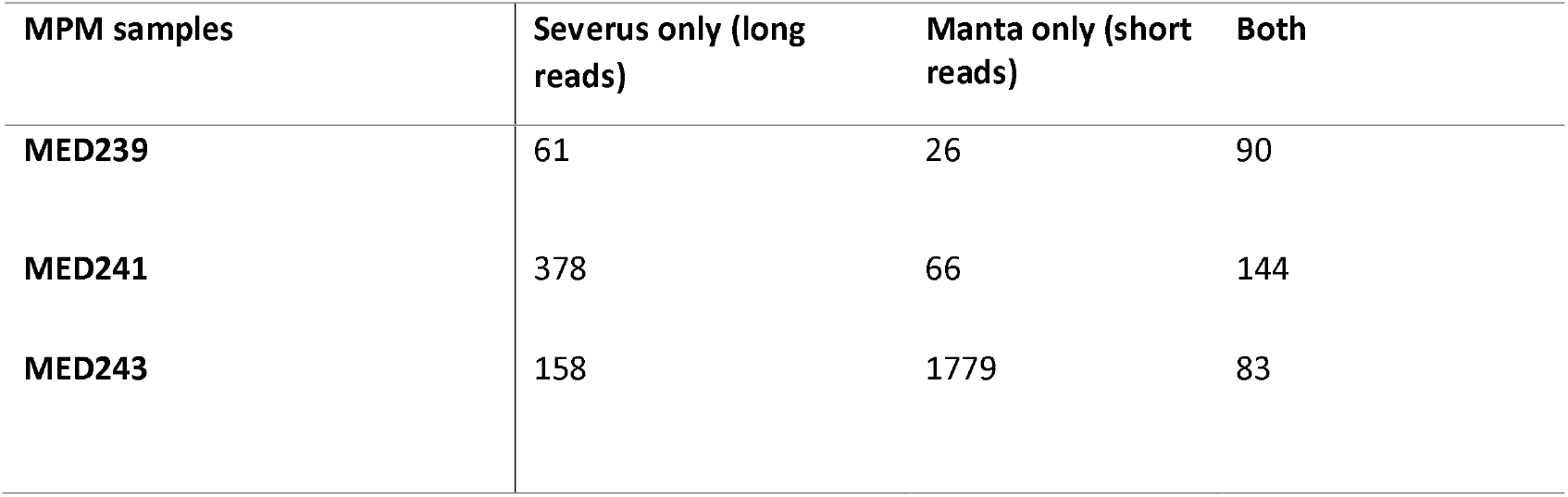
Number of somatic SVs identified using two callers from long-reads and short-reads.

| MPM samples | Severus only (long reads) | Manta only (short reads) | Both |
| --- | --- | --- | --- |
| MED239 | 61 | 26 | 90 |
| MED241 | 378 | 66 | 144 |
| MED243 | 158 | 1779 | 83 |

To evaluate the biological significance of these novel SVs, breakpoint of long reads specific SVs were intersect with the Catalog of Somatic Mutations in Cancer (COSMICS) dataset [39] from https://cancer.sanger.ac.uk/cosmic. We assessed whether SVs breakpoints occurred within the genomic coordinates of known cancer-associated genes. This analysis identified twelve genes in which SV breakpoints were located with COSMIC-annotated regions, including *LRP1B, BIRC6, EXT1, NBEA, GPC5, LPP, ROBO2, JAZF1, CDH1, FLCN, SPECC1, EPHA7* (S10.A Table). In addition, SV breakpoints were identified in 116 genes not annotated in COSMIC. To evaluate their potential relevance, these genes were assessed against existing literature review, revealing several candidates – including *GPC6, PRKN, ENTR1, SH3RF3, ANTXR1, PEX5L, SULT1A2, WEE1* – that have potential biomarker gene or potential tumour suppressor gene in various type of cancers (S10.B Table) [40, 41, 42, 43, 44, 45, 46, 47, 48, 49].

To confirm the accuracy of SV predictions, selected breakpoint events were validated using PCR amplification and followed by Sanger sequencing. Three primer pairs were designed to target novel SVs affecting *WEE1, PRKN*, and *GPC6*. These experiments, confirmed both breakpoint coordinates and rearrangement types (S11A-B Figures).

We next integrated SV calls from both long-read and short-read sequencing with SNV data to identify the double hit events in known MPM driver genes. Double hits were observed in *NF2, LATS2, BAP1, RB1, LATS1* (Table 5), highlighting the contribution of SV to tumour suppressor gene inactivation in MPM. Independent SV detection using Severus and Manta confirmed the second hit structural variant affecting of several of these genes such as *LATS1, NF2*, and *BAP1*, which were further validated by PCR and Sanger sequencing (S12 Figure).

**Table 5:**
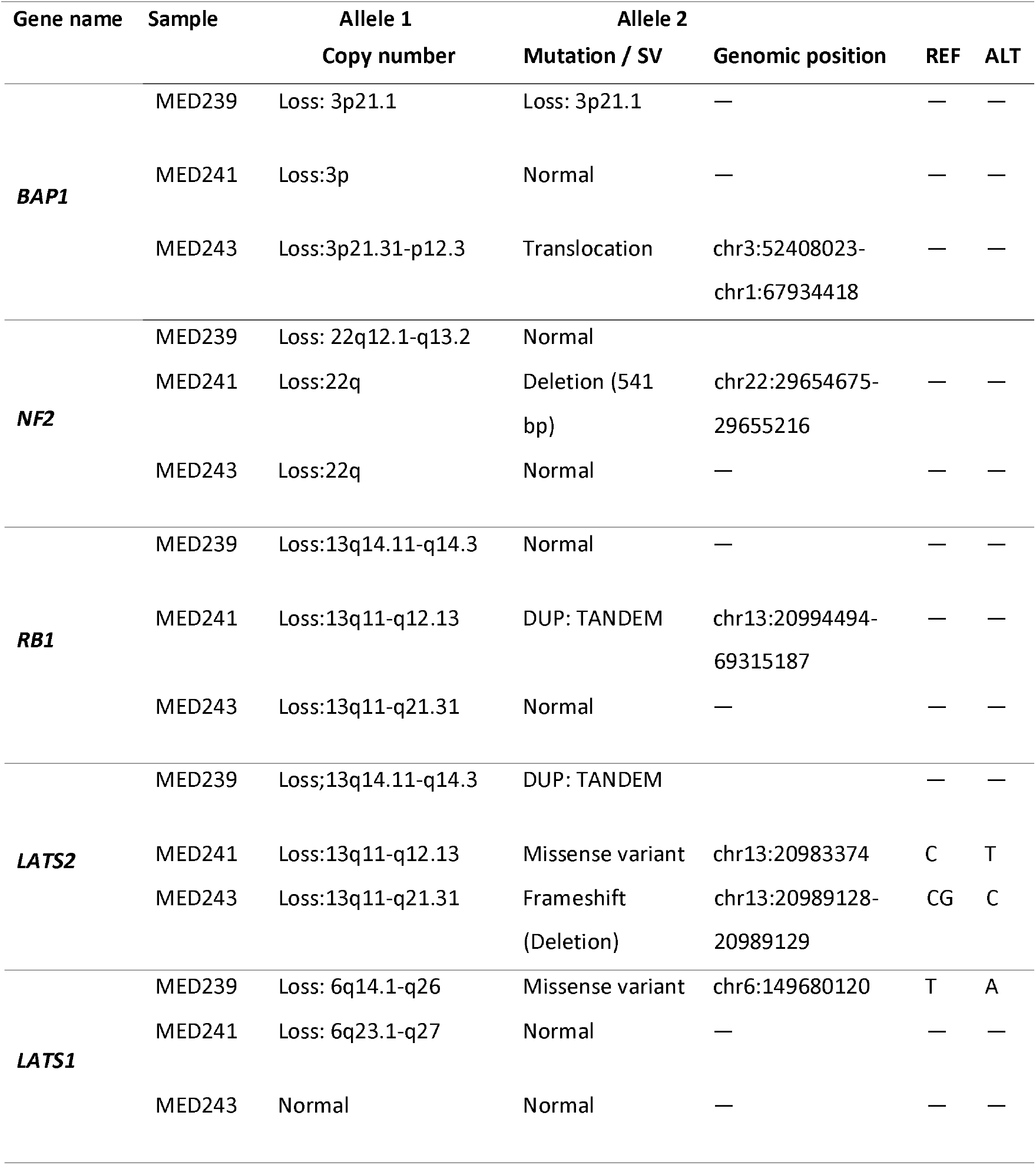
Double mutational hits of MPM driver genes.

Together, these findings indicate that structural variation represents a major driver of genomic and regulatory disruption in MPM, extending beyond copy number alterations alone.

### Complex structural rearrangements are identified only by long read sequencing

We used our long-read sequencing data to resolve structural rearrangements involving multiple chromosomes and SVs types. In MED239, complex SV clusters involved interlinked rearrangement across multiple chromosomes, including region affecting tumour suppressor genes such as *RB1* and *LATS2* (Figure 2, S13.A). In MED241, large-scale of rearrangements displayed features consistent with chromothripsis-like events, including oscillating copy numbers events (Figure S13.B). In MED243, extensive with rearrangements were observed on chromosome 16, affecting genes such as *CREBBP, CYLD*, and *PALB2* without corresponding copy number changes (Figure S13.C).

**Figure 2.**
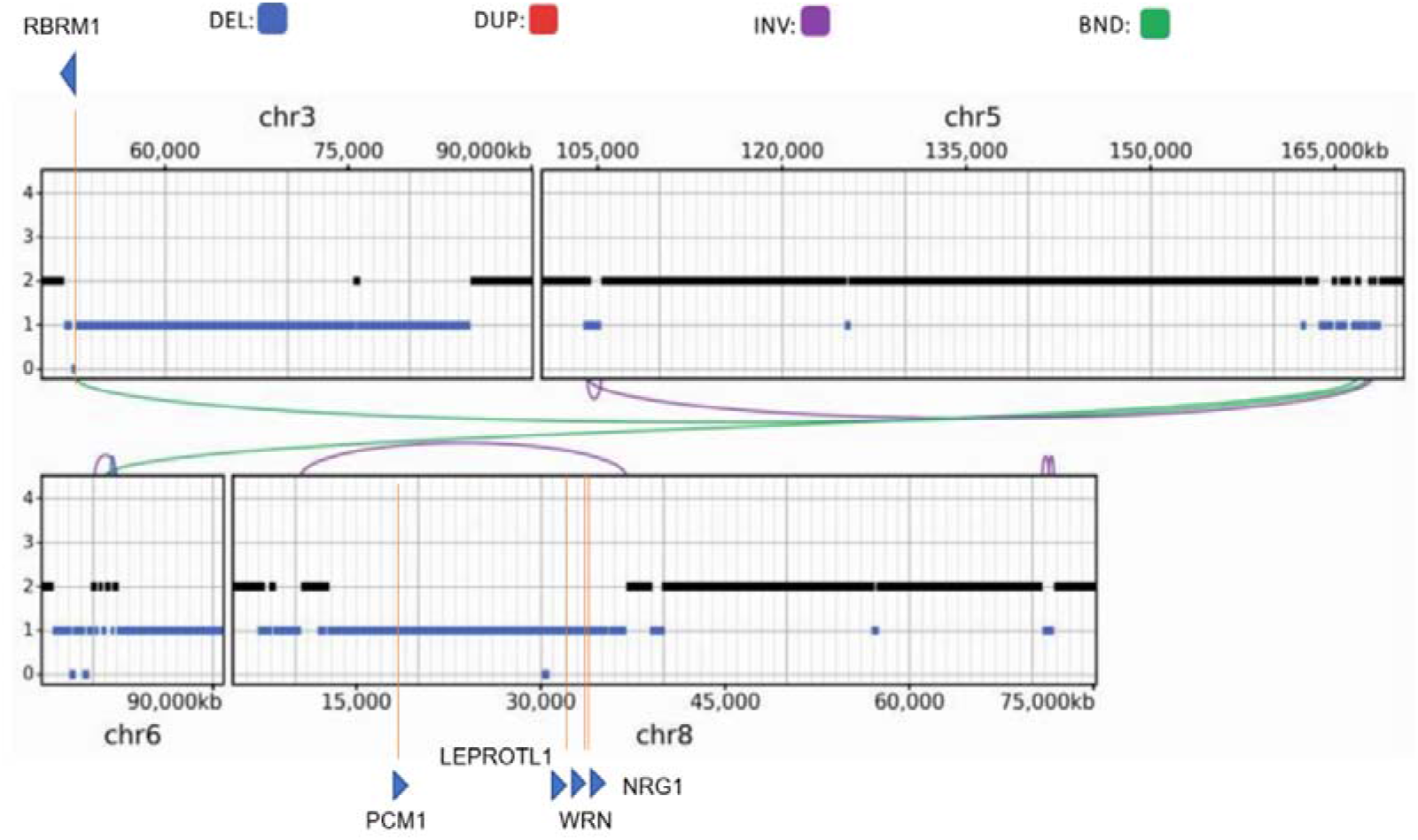
Copy Number and Structural Variation Profile of Cluster severus_1 among Chromosomes 3, 5, 6, and 8 in MED239: Shown is the copy number variation detected by CNVkit from short-read WGS and structural rearrangements detected by Severus from long read WGS in cluster Severus 0, visualised across chromosomes 3, 5, 6, and 8. The Y-axis represents copy number states, while coloured links indicate different types of structural variants: deletions (blue), duplications (red), inversions (purple), and breakends/translocations (green). Genes affected by these structural variations overlap with several COSMIC cancer-related genes highlighted in orange, which were *PBRM1, LEPROTLI, NRG1, PCM1*, and *WRN*. Notably, *PBRM1* on chr3 and *WRN/NRG1* on chr8 are disrupted in regions showing copy number loss.

These events formed clustered patterns of genomic instability consistent with chromoplexy and chromothripsis-like processes occurring extensively during the development of MPM. These findings indicate that complex structural rearrangements represent a major and previously underappreciated feature of the MPM genome.

### Functional and transcriptional impact of novel SVs

To assess the transcriptional impact of genomic alteration, we first performed a genome-wide analysis integrating gene expression (TPM) with copy number (CN) profiles derived from short-read WGS across three MPM tumour samples.

At the global level, comparison of genes with heterozygous loss (CN = 1) to copy-neutral genes (CN = 2) revealed that CN loss is generally associated with reduced gene expression, consistent with a gene dosage effect. This relationship was further supported by a positive, albeit modest, correlation between CN and gene expression (Spearman’s *rho* = 0.13, and 0.07 for MED239, MED241, and MED243 respectively; *p* < 2.2 x 10^-16^ ). However, the relatively weak correlations indicate that copy number alone explains only a portion of the observed transcriptional variability.

Having established this genome-wide baseline, we next investigated whether novel SVs identified by long-read WGS contribute to gene expression change beyond the copy number effects. To this end, SVs data were integrated with gene expression and CN profiles at the gene level.

Among the genes affected by novel SVs, several showed disrupted expression patterns. For example, *WEE1* affected by the large complex inversion in MED241 and in MED243 was affected by a tandem duplication spanning its promoter region and showed markedly increased expression, consistent with potential gene activation despite normal copy number (CN=2) (Table S14). Similarly, GPC6 in MED241, although copy-neutral (CN=2), exhibited upregulated expression coinciding with a novel duplication. In contrast, *PRKN* showed consistently low expression across all samples, despite variable copy number status and multiple SVs overlapping *PRKN* – some of which were detected only through long-read WGS, while others were also identified by short-read WGS. Other SV-affected genes such as *ENTR1, EPHA7*, and *CDH1* did not exhibit significant expression changes (Table S14).

Collectively, these findings demonstrate that while the genome-wide transcriptional patterns are partly explained by copy number variation, additional regulatory complexity exists. Novel SVs may contribute to gene specific expression change; however, given the limited sample size and variability observed these effects remain difficult to distinguish from background transcriptional noise.

### Clinical relevance of genes affected by SVs detected by long-read sequencing

Because SVs can affect gene expression, we measured the relationship between gene expression and MPM survival rates in order to indirectly assess the potential clinical relevance of SVs. We evaluated both of genes overlapping the COSMIC database and additional candidates previously reported as potential cancer biomarkers. Using the TCGA-MESO cohort, patients were stratified into high and low expression groups by median expression per gene, and survival differences were assessed using the log-rank test with Benjamini-Hochberg correction for p-values.

Four genes (*WEE1, GPC6, PRKN*, and *ENTR1*) showed statistically significant associations with overall survival (adjusted p < 0.05; S15 Table). *WEE1* had the strongest association with poor prognosis; patients in the high-expression group exhibited significantly shorter survival, particularly within the first two years (Figure 3). This is consistent with the known role of *WEE1* in cell cycle regulation and resistance to DNA damaging therapies, supporting its potential as a therapeutic target. Similarly, overexpression of *GPC6* correlated with worse outcomes (Figure 3), suggesting involvement in tumour progression, possibly via Wnt signaling.

**Figure 3:**
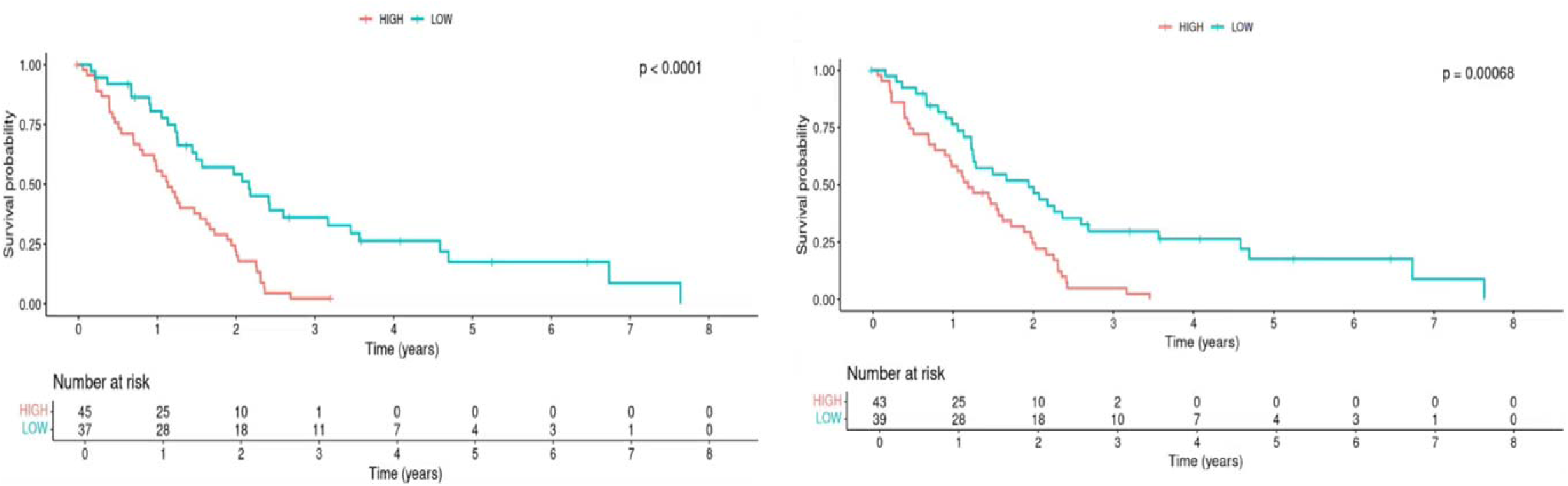
Kaplan–Meier survival analysis based on gene expression of WEE1 and GPC6 in mesothelioma patients from TCGA data. Patients were stratified into high (red) and low (blue) expression groups based on the median expression level for each gene. Each panel shows overall survival (OS) probability over time (years). The tables beneath each plot show the number of patients at risk at each time point. The left plot represents the survival time based on gene expression of WEE1 in mesothelioma patients. The right plot represents the survival time based on gene expression of GPC6 in mesothelioma patients.

### Analysis of methylation

To characterise the epigenomic landscape of MPM tumours, we analysed genome-wide CpG methylation profiles using long read sequencing data. Genome-wide methylation patterns were used to estimate cellular composition through methylation-based deconvolution. This analysis revealed substantial contributions from non-tumour cell population, including fibroblast (13-35%) and immune cells (25-61%) with corresponding population variability in tumour purity (S16 Table).

We next examined promoter-specific patterns. Promoter regions were defined as ±1 kb from annotated transcription start sites (TSS) and extracted using the methylkit. Analysis of promoter methylation showed a predominance of highly methylated states across all MPM samples. Consistent with this, the distribution of average promoter methylation showed a characteristic bimodal pattern, with enrichment of highly methylated promoters and no substantial global differences between samples were (Figure 4), indicating a broadly consistent promoter methylation landscape across MPM.

**Figure 4.**
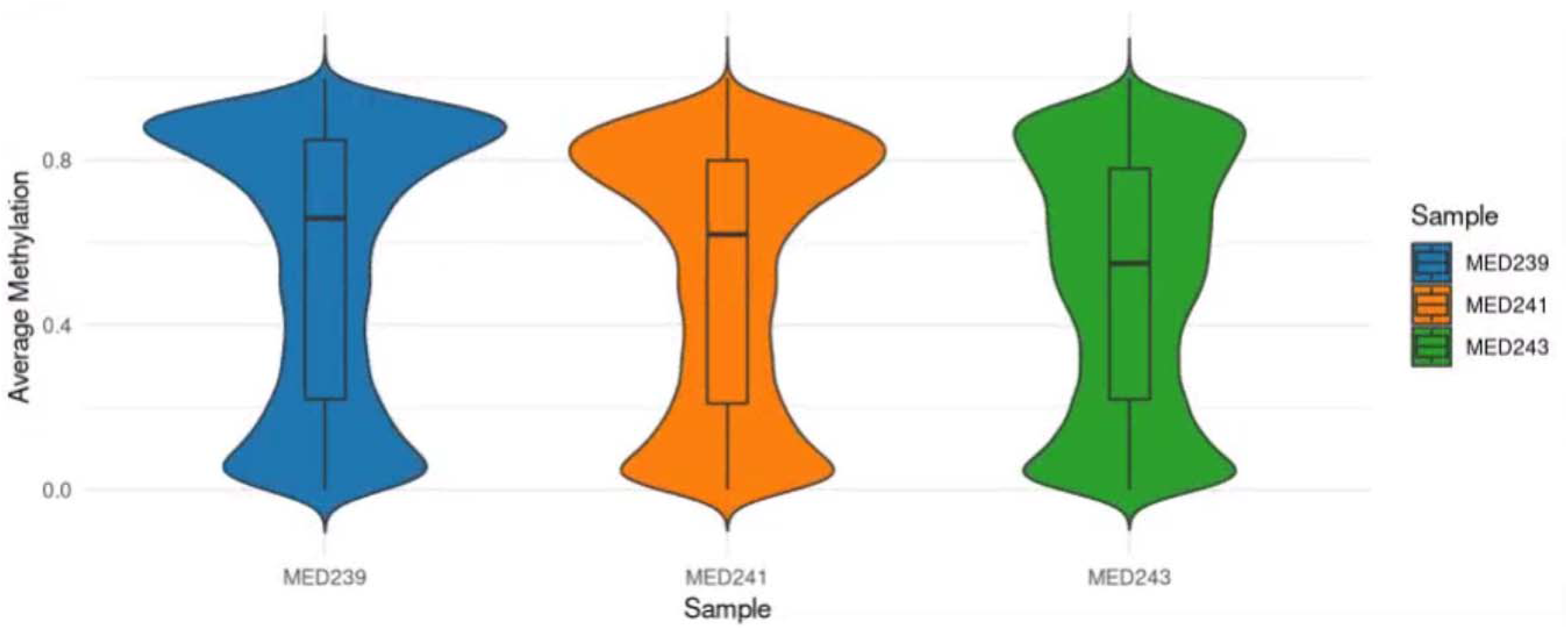
Distribution of average promoter methylation level across MPM tumour samples. Violin plots showing the distribution of average promoter methylation level for MED239, MED241, and MED243. The width of each violin represents the density of value at a given methylation level. Embedded boxplots indicate the median (centeral line), interquartile range (box), and range (whishers). Overall, similar methylation distributions were observed across the three sample, woth no substantial global differences.

Finally, we integrated methylation and structural variation, focusing on two for genes WEE1 and GPC6 affected by long-read-specific SVs. In two cases, WEE1 and GPC6 coincided with reduced promoter methylation and increased gene expression despite the absence of copy number alterations directly affecting these genes (Table 6). These observations suggest a potential link between structural variation, epigenetic modification, and transcriptional regulation. However, as this analysis was limited to a small number of genes, the findings should be interpreted with caution.

**Table 6.** Promoter differential methylation.

| Chromosome | start | end | strand | p value | q value | differential methylation | Gene name |
| --- | --- | --- | --- | --- | --- | --- | --- |
| chr11 | 9572669 | 9574669 | + | 1.01E-29 | 1.40E-28 | -1.9426 | *WEE1* |
| chr11 | 9573686 | 9575686 | + | 0.7007 | 0.313215 | 0.03361 | *WEE1* |
| chr11 | 9574492 | 9576492 | + | 9.90E-20 | 7.08E-19 | -4.0866 | *WEE1* |
| chr11 | 9577027 | 9579027 | + | 2.43E-11 | 7.77E-11 | -16.344 | *WEE1* |
| chr13 | 93225806 | 93227806 | + | 0.002032 | 0.00265567 | -2.08656 | *GPC6* |
| chr13 | 94113122 | 94115122 | + | 1.70E-08 | 5.71E-08 | -13.2662 | *GPC6* |
| chr13 | 93857473 | 93859473 | - | 0.13383 | 0.09960276 | -3.44256 | *GPC6* |

Together, these results support a model in which structural variation may contribute to tumour progression not only through direct genomic disruption but also by reshaping the epigenetic landscape, leading to transcriptional dysregulation independent of gene dosage. Although limited to a subset of genes, these findings provide preliminary evidence linking structural variation to epigenetic and transcriptional regulation in MPM.

## Discussion

A comprehensive catalogue of genetic variation in tumours is critical in understanding their origin, development and treatment vulnerabilities [50]. In particular, studies across many different types of cancer have shown that SVs are frequent driver mutations, yet routine comprehensive discovery of these SVs remains challenging, with the result that they are an understudied class of driver mutation in cancer genomics [51]. Recent studies show that a full appreciation of SVs facilitate different clinical applications, including early detection and prognosis [52]. In MPM, the role of large-scale SVs is appreciated [53], and it is known that large-scale chromosomal arm loss, rather than single nucleotide changes, drive the early evolution of the disease [53]. Furthermore, gene fusions in MPM have been identified [54], further supporting the critical role of SVs in the disease.

In this study we used long read sequencing to explore the hidden SV landscape of the MPM genome. We found evidence of SVs affecting known MPM driver genes, sometimes comprising a second-hit event. We also find evidence of complex events involving multiple chromosomes reflecting a history of chromoplexy in the tumour. Our data support previous studies [55, 56] which suggest chromoplexy is a major feature of MPM.

An advantage of long read sequencing using Oxford Nanopore technology is that CPG methylation can be directly detected by the nanopore itself, requiring no chemical bisulphite treatment. Because the interplay of structural variation and epigenetic variation in cancer [57, 58], we used this feature of nanopore sequencing to study the epigenome of MPM in the context of structural variation. We focused on promoter methylation, because methylation of CpG islands upstream of transcription start sites is most clearly linked to gene expression, and found very similar patterns of methylation across the tumours analysed, consistent with previous analysis of methylation array data [59]. However, we found two specific examples where two oncogenes (WEE1, GPC6) next to large scale SVs, but with unaltered copy number, showed promoter hypomethylation and increased expression levels. DNA methylation has been used as the molecular basis of liquid-biopsy approach for other cancers [60], and this represents a promising avenue for developing an early-detection test for MPM. Furthermore, the possibility of directly modifying DNA methylation suggests possible future therapeutic approaches for MPM, emphasizing the need for routine collection of rich and robust data on the MPM epigenome [61, 62]

Beyond gene-specific examples, our results suggest that SV may contributes more broadly to transcriptional dysregulation in MPM. Genome-wide analysis demonstrated that copy number alterations, which often arise from large-scale structural events, are associated with corresponding changes in gene expression, with copy number loss generally linked to reduce expression level, consist with gene dosage effects observed in MPM [5, 10]. However, the relatively modest correlation observed between copy number and expression indicates that additional mechanisms contribute to transcriptional variability.

Structural variants may influence gene expression through multiple mechanisms, including disruption of gene bodies, alteration of regulatory elements, and reorganization of chromatin architecture [63]. In particular, SVs can reposition enhancers or promoters, leading to aberrant gene activation or repression independent of copy number changes, a phenomenon often referred to as enhancer hijacking [64]. Complex rearrangements such as chromoplexy, which have been described in cancer genomes, have the potential to simultaneously affect multiple loci and drive coordinated transcriptional changes [65].

While only a limited number of gene-specific examples were identified in this study, these observations are consistent with a broader role for SVs in shaping the transcriptional landscape of MPM. Together, these finding support the view that structural variation may contributes to gene expression not only through direct gene disruption but also via complex regulatory mechanisms.

A notable finding of this study is the limited overlap in SNV detection between short-read WGS and long-reads WGS approaches. Across all samples, only a small subset of variants was consistently identified by both platforms, particularly for functionally relevant variants such as missense and stop-gained mutations. This low concordance highlights current challenges in SNV detection using long reads technologies. Several factors likely contribution to this discrepancy. Long-read sequencing is characterized by higher base-level error rates compared to short-read platforms, which can impact the accuracy of SNV calling. In addition, differences in variant calling algorithms, including NanoCaller and ClairS, result in variable sensitivity and specificity, as reflected by the differing numbers of SNVs detected across caller. Sequencing depth is also a critical factor, as insufficient coverage may reduce confidence in variant detection and contribute to inconsistencies between platforms.

At present, short-read WGS remains the standard approach for somatic SNV detection in both research and clinical settings, owing to its higher base accuracy and well-established analytical pipelines. In contrast, while long-read sequencing offering clear advantage for detection SVs and resolve complex genomics regions, its application for SNV detection is still evolving requires further methodological development and optimization. Similar limitations in long-read SNV calling and platform concordance have been reported in previous studies, including in cancer genome analysis using both sequencing technologies [66].

The main limitation of this study is the small sample size of three patients, which restricts our ability to directly associate particular SVs with key clinical parameters, including survival and response to treatment. It also prevents us identifying frequent SVs that might indicate novel driver genes not previously identified by recurrent SNV analysis. However, this study provides a basis and justification for Nanopore long-read sequencing on larger MPM cohorts because of the large yield of SVs undetected by short-read sequencing, and the full profiling of the epigenome, both of which are likely to have major clinical relevance for this disease.

## Data Availability

All data produced in the present study are available upon request to the authors, subject to review for compliance with data protection legislation.

## Acknowledgements

This research used the ALICE High Performance Computing facility at the University of Leicester. This work was supported in part by a British Lung Foundation-Mesothelioma UK grant MESOUK17-8.

MHA is supported by the Ministry of Education, under the program of Custodian of the Two Holy Mosques, and the Royal Embassy of Saudi Arabia Cultural Bureau.

## Author contributions

MHA – Conceptualisation, Data Curation, Formal Analysis, Investigation, Methodology, Visualisation, Validation, Writing – original draft

CP – Data curation, Resources

JD, AB, KK. Resources

DF – Funding acquisition, Resources

EJH – Conceptualisation, Funding acquisition, Resources, Supervision, Writing-review and editing

## Notes

### Competing Interest Statement

The authors have declared no competing interest.

### Author Declarations

NHS research ethics committees references 14/LO/1527 (London/Fulham) and 14/EM/1159 (East Midlands/Leicester South) gave ethical approval for this work.

