## Supplementary Material for "Long-read Oxford Nanopore sequencing reveals complex rearrangements and regulatory disruption in malignant pleural mesothelioma"

**S1 Table** **Summary of the PCR primers design (5' to 3') used to validate SVs in patient samples.**

|  | SV event | pair sequence (5' to 3') | PCR product size (bp) | |
| --- | --- | --- | --- | --- |
| MED243 | 11:9560349-9576225  DUP | F-ACTCCGGATTCTTTGTTGCT | | 243 |
|  |  | R-AAAATGTGGGGAGACAAAGTTAA | |  |
| MED241 | 13:93767822:93848124  DUP | F-TGCTTGAAGATGAGTTAACGTT | | 286 |
|  |  | R-CTGAAGAGCTGACTTTGGGC | |  |

**S2 Table Summary of clinical information.**

| Sample | Sex | Age | Ethnicity | Asbestos exposure | Smoking status | Side | Histology |
| --- | --- | --- | --- | --- | --- | --- | --- |
| MED239 | Male | 60-69 | White British | Yes | Ex-smoker | R | Epithelioid |
| MED241 | Male | 70-79 | White British | No | Ex-smoker | R | Epithelioid |
| MED243 | Male | 60-69 | Black British | Unknown | Unknown | L | Epithelioid |

**S3.A Table** **Summary of sequencing output after basecalling with Guppy for the tumour and non-tumour blood samples.**

|  | **MED239** | | **MED241** | | **MED243** | |
| --- | --- | --- | --- | --- | --- | --- |
|  | **Tumour** | **Blood** | **Tumour** | **Blood** | **Tumour** | **Blood** |
| **N50 (kb)** | 20 | 18 | 24 | 21 | 28 | 25 |
| **Total bases (Gb)** | 119.1 | 144.3 | 130.8 | 120.6 | 122 | 131.6 |
| **Estimated coverage (fold)** | 38 | 46 | 41 | 38 | 39 | 42 |

**S3.B Table** **Quality Assessment of Sequencing Reads (%) After Basecalling with Guppy:** Q5 thresholds indicated that the reads with a >1 in 3 error rate were filtered out.

|  | MED239 | | MED241 | | MED243 | |
| --- | --- | --- | --- | --- | --- | --- |
|  | **Tumour** | **Blood** | **Tumour** | **Blood** | **Tumour** | **Blood** |
| **> Q5** | 100.0 | 100.0 | 100.0 | 100.0 | 100.0 | 100.0 |
| **> Q7** | 100.0 | 100.0 | 99.3 | 99.4 | 99.7 | 99.6 |
| **> Q10** | 95.9 | 96.6 | 95.1 | 95.3 | 96.4 | 96.5 |
| **> Q12** | 92.1 | 93.4 | 92.0 | 91.3 | 94.1 | 93.7 |
| **> Q15** | 80.0 | 85.3 | 81.1 | 78.2 | 85.8 | 84.8 |

|  | **MED239** | | **MED241** | | **MED243** | |
| --- | --- | --- | --- | --- | --- | --- |
|  | **Tumour** | **Blood** | **Tumour** | **Blood** | **Tumour** | **Blood** |
| 1 | 580 | 601 | 482 | 256 | 248 | 687 |
| 2 | 423 | 597 | 229 | 246 | 220 | 393 |
| 3 | 314 | 537 | 203 | 195 | 216 | 302 |
| 4 | 184 | 510 | 191 | 191 | 215 | 242 |
| 5 | 182 | 502 | 191 | 186 | 208 | 233 |

**S3.C Table** **Top Five Longest Sequencing Reads (kb) for Tumour and Blood Samples.**

**S3.D Table	Summary of mapped reads After mapping with minimap2 for Tumour and Blood Samples.**

|  | **MED239** | | | **MED241** | | **MED243** | | |
| --- | --- | --- | --- | --- | --- | --- | --- | --- |
|  | **Tumour** | | **Blood** | **Tumour** | **Blood** | **Tumour** | | **Blood** |
| **N50 (kb)** | 25.5 | 19.6 | | 25.5 | 22.2 | 29.2 | 26.1 | |
| **Total of bases aligned (gb)** | 118.8 | 144.1 | | 130.5 | 120.2 | 122 | 131.5 | |
| **Estimated coverage (fold)** | 38.3 | 46.4 | | 42.09 | 38.7 | 39.3 | 42.4 | |
| **Percentage of reads Aligned** | 99.78 | 99.82 | | 99.42 | 99.43 | 99.67 | 99.67 | |

**S4 Table** **Summary of missense variants and stop-gained mutations identified in at least two MPM tumour samples:** this analysis filtered based on identical genomic positions (same SNV) or shared mutant genes (different positions).

| Found in at least 2 samples | Missense variants | Stop gained |
| --- | --- | --- |
| **Same SNV** | 23 | 0 |
| **Same SNV overlap with COSMIC** | 7 | 0 |
| **Mutated gene** | 353 | 3 |
| **Mutated gene overlap with COSMIC** | 127 | 0 |

**S5 Table summary of somatic copy number alteration in three patient sample:** this table illustrate the total number of copy number of segmentations, the total of copy number loss either homozygous or heterozygous loss represented in deletion column, and the copy number gained represented in amplification column, the total length of both loss and gain represented in Total Length (Mb) column, and their average length represented in average length (kb) column.

| Samples | Total number of segmentations CN found | Total CNAs | Deletion | Amplification | Total Length (Mb) | Average length (kb) |
| --- | --- | --- | --- | --- | --- | --- |
| MED239 | 1537 | 605 | 502 | 103 | 442.9 | 882.3 |
| MED241 | 1171 | 643 | 564 | 79 | 602.4 | 1103.3 |
| MED243 | 905 | 485 | 399 | 86 | 324.7 | 813.9 |

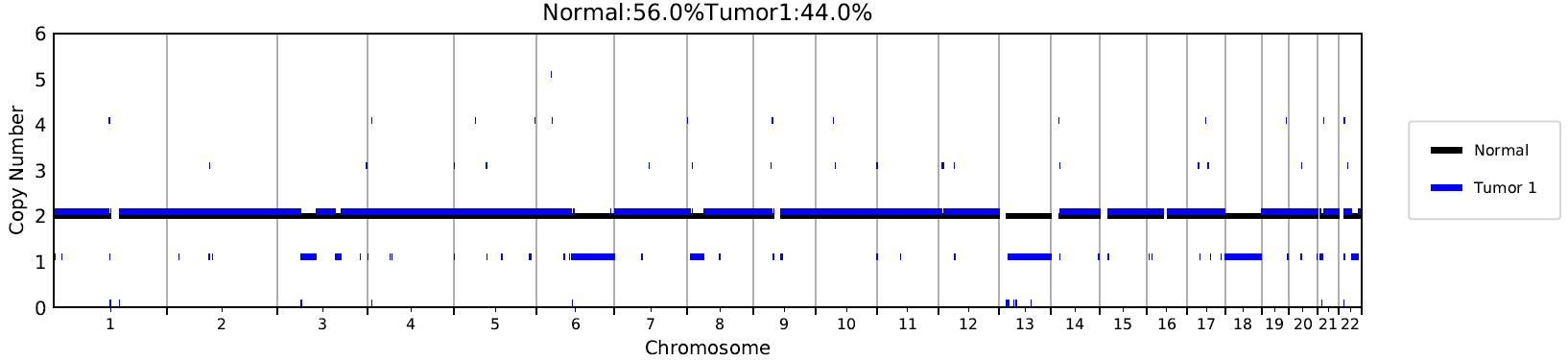

**S 6.A Figure** **Whole-genome profiles of CNAs by CNVkit in MED239 sample:** the Y-axis indicates the absolute copy number and X-axis indicates the chromosomes from chr1 to chr22. The bule line represent the copy number of the tumour sample, while the black line represents the copy number of the matched normal samples.

**S 6.B Figure Whole-genome profiles of CNAs by CNVkit in MED241 sample:** the Y-axis indicates the absolute copy number and X-axis indicates the chromosomes from chr1 to chr22. The bule line represent the copy number of the tumour sample, while the black line represents the copy number of the matched normal samples.

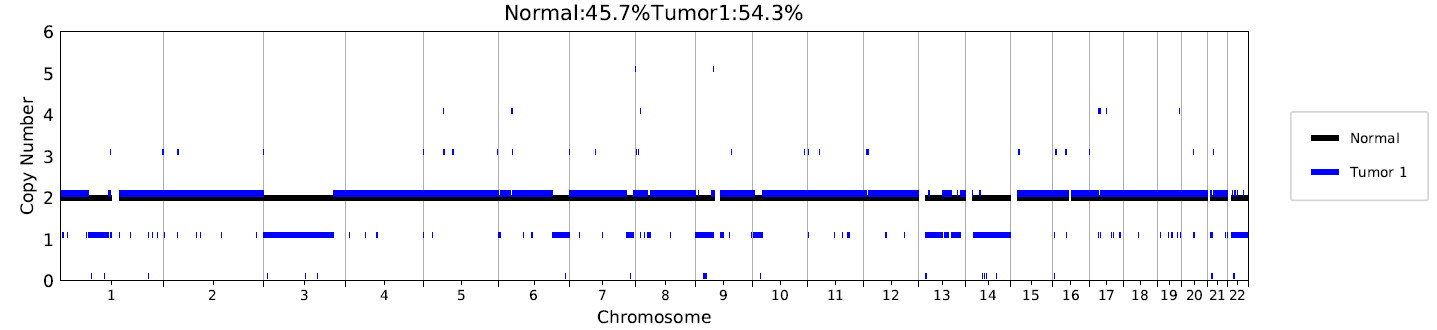

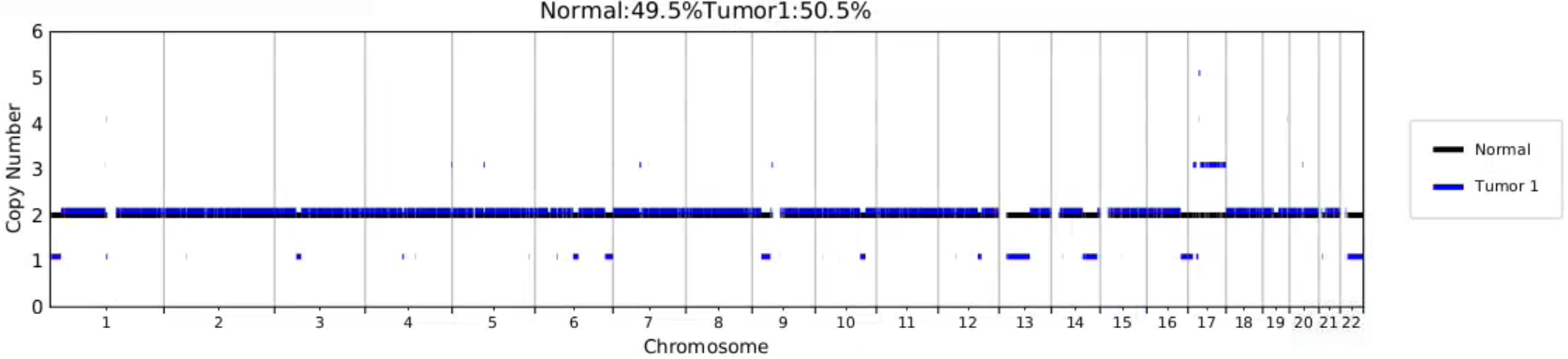

**S 6.C Figure Whole-genome profiles of CNAs by CNVkit in MED243 sample:** the Y-axis indicates the absolute copy number and X-axis indicates the chromosomes from chr1 to chr22. The bule line represent the copy number of the tumour sample, while the black line represents the copy number of the matched normal samples.

**S7 Table Somatic Structural Variations Identified in MPM Patients Using Manta:** The data are categorised by patient samples (MED239, MED241, and MED243) and different classes of SVs: DELs (the loss of a DNA segment), translocation (BNDs), structural rearrangements where a breakpoint is identified), DUPs (the duplication of a DNA segment), INVs (a segment of DNA reversed within the genome), and insertions (the addition of new DNA sequences).

|  | **MED239** | **MED241** | **MED243** |
| --- | --- | --- | --- |
| **DEL** | 27 | 93 | 50 |
| **BND** | 58 | 60 | 1,740 |
| **DUP** | 10 | 17 | 15 |
| **INV** | 17 | 41 | 56 |
| **INS** | 0 | 0 | 0 |
| **total** | 112 | 211 | 1,861 |

**S8.A Table Somatic** **Structural Variations Identified in MPM Patients:** This table summarises the number and types of somatic structural variations (SVs) detected in three patient samples (MED239, MED241, and MED243) using Sniffles; DEL (the loss of a DNA segment), translocation (BND), structural rearrangements where a breakpoint is identified), DUP (the duplication of a DNA segment), INV (a segment of DNA reversed within the genome), and insertions (the addition of new DNA sequences).

|  | MED239 | MED241 | MED243 |
| --- | --- | --- | --- |
| DEL | 52 | 82 | 248 |
| INS | 107 | 109 | 1389 |
| BND | 9 | 3 | 11 |
| DUP | 3 | 1 | 5 |
| INV | 5 | 2 | 15 |
| total | 176 | 197 | 1,688 |

**S8.B Table Somatic Structural Variations Identified in MPM Patients:** This table summarises the number and types of somatic structural variations (SVs) detected in three patient samples (MED239, MED241, and MED243) using Severus; DEL (the loss of a DNA segment), translocation (BND), structural rearrangements where a breakpoint is identified), DUP (the duplication of a DNA segment), INV (a segment of DNA reversed within the genome), and insertions (the addition of new DNA sequences).

|  | **MED239** | **MED241** | **MED243** |
| --- | --- | --- | --- |
| **DEL** | 42 | 98 | 91 |
| **INS** | 32 | 26 | 38 |
| **BND** | 54 | 60 | 58 |
| **DUP** | 6 | 8 | 9 |
| **INV** | 18 | 38 | 47 |
| **total** | 151 | 230 | 243 |

**S8.C Table Summary of the overlap of somatic SVs between the by two SV callers using Nanopore sequencing across MPM patient samples**.

| **Samples/SV caller** | Severus | Sniffles | Both |
| --- | --- | --- | --- |
| **MED239** | 123 | 146 | 27 |
| **MED241** | 212 | 186 | 10 |
| **MED243** | 170 | 1494 | 70 |

**S9 Table** **Novel Somatic Structural Variations Identified in MPM Patients:** It summarises the number and types of novel somatic SVs detected in three patient samples (MED239, MED241, and MED243) using only Severus from long read WGS. The subsequent sections detail the counts of specific SV classes, including DEL, INS, BND, DUP, and INV, as detected by Severus.

|  | MED239 | MED241 | MED243 |
| --- | --- | --- | --- |
| DEL | 19 | 30 | 54 |
| BND | 2 | 8 | 28 |
| DUP | 1 | 3 | 5 |
| INV | 7 | 14 | 33 |
| INS | 32 | 23 | 38 |
| Total | 61 | 78 | 156 |

**S10.A Table Identification of somatic SVs affecting cancer genes (COSMIC database)**.

| **Chromosome** | **Start** | **End** | **Length (bp)** | **SV type** | **Gene name** | **Sample** |
| --- | --- | --- | --- | --- | --- | --- |
| 2 | 32461063 | 32461064 | 186 | INS | *BIRC6* | MED241 |
| 2 | 141176486 | 141187861 | 11375 | DEL | *LRP1B* | MED241 |
| 3 | 35110024 | 35111263 | 626 | DEL | *NBEA* | MED241 |
| 3 | 77169869 | 77169870 | 0 | TRA | *ROBO2* | MED243 |
| 3 | 188607485 | 188648635 | 41150 | DEL | *LPP* | MED243 |
| 6 | 83262522 | 94538205 | 11275683 | DEL | *EPHA7* | MED243 |
| 7 | 28081536 | 28081537 | 0 | TRA | *JAZF1* | MED243 |
| 8 | 117861499 | 117863098 | 1599 | INV | *EXT1* | MED241 |
| 13 | 35099198 | 35100240 | 335 | DEL | *NBEA* | MED241 |
| 13 | 35101065 | 35102299 | 1234 | DEL | *NBEA* | MED241 |
| 16 | 67810595 | 72737783 | 4927188 | DUP | *CDH1* | MED243 |
| 17 | 17195434 | 20425437 | 3230003 | DEL | *FLCN* | MED243 |
| 17 | 17195434 | 20425437 | 3230003 | DEL | *SPECC1* | MED243 |

| Chromosome | Start | End | Region | Affecting Coding region | | Length (bp) | SV Type | Gene name | Samples |
| --- | --- | --- | --- | --- | --- | --- | --- | --- | --- |
| 2 | 69096288 | 69096289 | intron | no | | 132 | INS | *ANTXR1* | MED239 |
| 2 | 69096288 | 69096289 | intron | no | | 55 | INS | *ANTXR1* | MED243 |
| 2 | 109199495 | 109199496 | intron | no | | 177 | INS | *SH3RF3* | MED239 |
| 2 | 109199495 | 109199496 | intron | no | | 928 | INS | *SH3RF3* | MED241 |
| 2 | 109199495 | 109199496 | intron | no | | 803 | INS | *SH3RF3* | MED243 |
| 3 | 179891202 | 179891203 | intron | no | | 840 | INS | *PEX5L* | MED243 |
| 6 | 155960356 | 169788723 | intron | exons | | 13828367 | DEL | *PRKN* | MED241 |
| 6 | 162010978 | 162030823 | intron-intron | | no | 19845 | INV | *PRKN* | MED241 |
| 9 | 136409423 | 136409424 | Intron | Promoter/Enhancer | | 99 | INS | *ENTR1* | MED241 |
| 11 | 3602648 | 71912867 | Intron | - | | 68310219 | INV | *WEE1* | MED242 |
| 11 | 9560349 | 9576225 | intron | promoter | | 15876 | DUP | *WEE1* | MED243 |
| 13 | 93767822 | 93848124 | intron-intron | | exon3 | 80302 | DUP | *GPC6* | MED241 |
| 13-22 | 93520820 | 16216680 | intron | no | | 0 | BND | *GPC6* | MED241 |
| 16 | 28591953 | 28591954 | exon | exon4 | | 1063 | INS | *SULT1A2* | MED243 |

**S10.B Table Identification of novel somatic SVs affected** **potential related cancer gene:** This table shows the novel SVs identified using only Severus from long reads WGS, these affected were previously literature as a potential biomarker gene or potential tumour suppressor gene in various type of cancers.

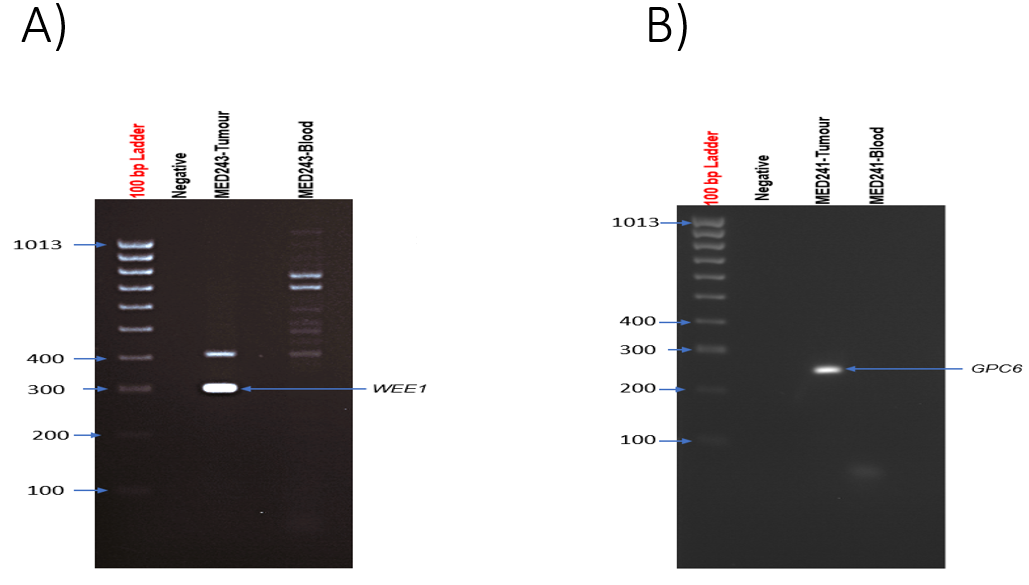

**S11.A Figure** **Validation of of novel SVs in MPM tumour by PCR:** Agarose gel electrophoresis showing PCR amplification product were present in tumour samples. A) 243 products flanking the breakpoint of tandem duplicated of WEE1 and B) 286 bp product flanking the breakpoint of tandem duplicated of GPC6.

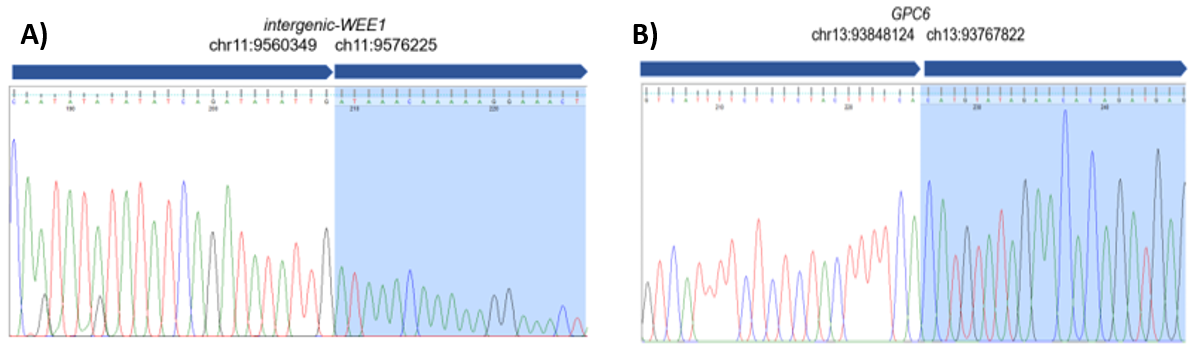

**S11.B Figure Validation of novel SVs in MPM tumour by Sanger sequencing:** A) Sanger sequencing traces confirming the 80 kb of tandem duplicated of GPC6 in sample MED241. B) Sanger sequencing traces confirming the the 15 kb of tandem duplicated of WEE1 in MED243.

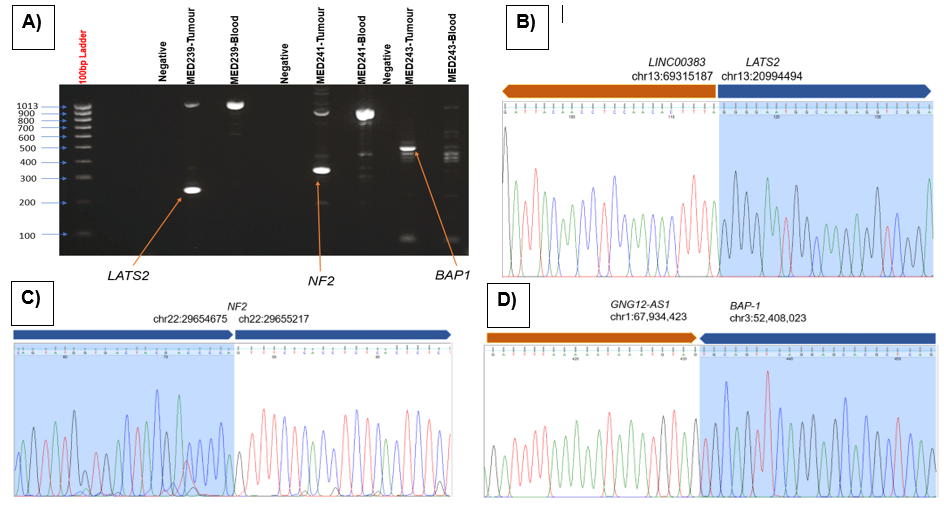

**S12 Figure Validation of double hit of three TSGs in MPM tumour by PCR and Sanger sequencing.** A) Agarose gel electrophoresis showing PCR amplification product were present in tumour samples; Negative means control with no genomic DNA, 245 bp products flanking the breakpoint of tandem duplicated of LATS2, 306 bp product flanking the breakpoint of 541bp deletion of NF2 (truncation mutant), and 455 bp product flanking the breakpoint of translocation of BAP1 to GNG12-AS1. B) Sanger sequencing traces confirming the beakpoint of tandem duplicated of LATS2 to. genomic DNA. C) Sanger sequencing traces confirming the deletion breakpoint of 541 bp deletion in sample MED241. D) Sanger sequencing traces confirming thebreak points of translocation of BAP1 to GNG12-AS1 in sample MED243.

**Table 13.A Summary of complex structural variation identified in MED239 using Severus from long reads WGS:** *hom =* *homozygous, het = Heterozygous, read support = number of the reads supporting this variant.*

| Sample | Cluster id | Strand A | | Breakpoint A | Strand B | | | | | | Breakpoint B | SVs Type details | | Read support | | | Genotype |
| --- | --- | --- | --- | --- | --- | --- | --- | --- | --- | --- | --- | --- | --- | --- | --- | --- | --- |
| MED239 | severus_0 | - | chr3:52677075 | | | - | | | chr5:167707772 | | | Complex inversion | | | 3 | | het |
|  | severus_0 | - | chr5:104147629 | | | - | | | chr5:105265736 | | | Complex inversion | | | 9 | | het |
|  | severus_0 | + | chr5:104184742 | | | + | | | chr5:168022821 | | | Complex inversion | | | 5 | | het |
|  | severus_0 | - | chr5:166838126 | | | - | | | chr6:81007568 | | | Inverted translocation | | | 5 | | hom |
|  | severus_0 | + | chr6:80133514 | | | + | | | chr6:81932467 | | | Inverted translocation | | | 9 | | het |
|  | severus_0 | + | chr6:81486010 | | | - | | | chr6:81798618 | | | Deletion | | | 10 | | hom |
|  | severus_0 | - | chr8:10469234 | | | - | | | chr8:36950137 | | | Complex inversion | | | 12 | | hom |
|  | severus_0 | + | chr8:37926374:INS:85 | | | | | |  | | | Insertion | | | 3 | | hom |
|  | severus_0 | + | chr8:70840897 | | | + | | | chr8:71372237 | | | Complex inversion | | | 6 | | het |
|  | severus_0 | - | chr8:71367686 | | | - | | | chr8:71766403 | | | Complex inversion | | | 6 | | het |
|  | severus_1 | + | chr13:19932416 | | | | - | chr13:69315208 | | | | | Deletion | | 11 | hom | |
|  | severus_1 | - | chr13:19932421 | | | | + | chr13:32101437 | | | | | Templated inserted | | 9 | hom | |
|  | severus_1 | - | chr13:21107179 | | | | + | chr13:70595047 | | | | | Templated inserted | | 8 | hom | |
|  | severus_1 | + | chr13:21113291 | | | - | | | | chr13:39554850 | | Deletion | | | 11 | | hom |
|  | severus_1 | + | chr13:31862430 | | | - | | | | chr13:69297068 | | Deletion | | | 10 | | het |
|  | severus_1 | - | chr13:31862441 | | | + | | | | chr13:39167716 | | Templated inserted | | | 10 | | het |
|  | severus_1 | + | chr13:31968333 | | | - | | | | chr13:31982163 | | Deletion | | | 11 | | het |
|  | severus_1 | - | chr13:32176860 | | | - | | | | chr13:39061181 | | Inversion | | | 7 | | het |

**Table 13.B Summary of complex structural variation identified in MED241 using Severus from long reads WGS:** hom = homozygous, het = Heterozygous, read support = number of the reads supporting this variant.

| Sample | Cluster id | Strand A | Breakpoint A | Strand B | Breakpoint B | SVs Type details | Read support | Genotype |
| --- | --- | --- | --- | --- | --- | --- | --- | --- |
| MED241 | severus_0 | - | chr11:83984628 | + | chr11:96567309 | Tandem duplicated | 4 | het |
|  | severus_0 | + | chr11:84052450 | + | chr11:99887070 | Inverted translocation | 4 | het |
|  | severus_0 | + | chr11:84574412 | - | chr11:96809434 | Deletion | 5 | hom |
|  | severus_0 | + | chr11:97511234 | - | chr13:64521454 | Inverted translocation | 5 | het |
|  | severus_0 | - | chr11:99443945 | - | chr7:4767076 | Templated inserted | 4 | het |
|  | severus_0 | - | chr11:99858245 | + | chr17:73546604 | translocation | 6 | het |
|  | severus_0 | - | chr11:99888047 | + | chr17:73635955 | translocation | 5 | het |
|  | severus_0 | - | chr11:100150518 | + | chr11:99978300 | Deletion | 4 | het |
|  | severus_0 | + | chr11:100242335 | - | chr11:84005960 | Templated inserted | 5 | hom |
|  | severus_0 | - | chr13:64366728 | - | chr13:71227416 | Complex inversion | 4 | hom |
|  | severus_0 | - | chr13:69995319 | + | chr13:71705663 | Templated inserted | 6 | hom |

***Table 13.C* Summary of complex structural variation identified in MED243 using Severus from long reads WGS:** *hom = homozygous, het = Heterozygous, read support = number of the reads supporting this variant.*

| Sample | Cluster id | Strand A | Breakpoint A | Strand B | Breakpoint B | SVs Type details | Read support | genotype |
| --- | --- | --- | --- | --- | --- | --- | --- | --- |
| MED243 | severus_0 | + | chr16:3078980 | - | chr16:67930422 | Deletion | 7 | hom |
|  | severus_0 | - | chr16:3079100 | - | chr16:67937944 | Complex inversion | 9 | het |
|  | severus_0 | + | chr16:3081294 | + | chr16:3089475 | Foldback inversion | 9 | het |
|  | severus_0 | - | chr16:3083968 | + | chr16:67823901 | Tandem duplicated | 5 | hom |
|  | severus_0 | + | chr16:3084139 | + | chr16:67934476 | Complex inversion | 5 | hom |
|  | severus_0 | - | chr16:3086645 | + | chr16:67827133 | Tandem duplicated | 7 | het |
|  | severus_0 | - | chr16:3090665 | + | chr16:67803452 | Templated inserted | 8 | het |
|  | severus_0 | - | chr16:67810595 | + | chr16:72737783 | Tandem duplicated | 7 | het |
|  | severus_0 | - | chr16:67826244 | - | chr1:23402320 | Complex inversion | 7 | het |
|  | severus_1 | - | chr3:132610989 | - | chr9:70898966 | Templated inserted | 6 | het |
|  | severus_1 | + | chr3:132610966 | + | chr9:71329676 | Templated inserted | 5 | het |
|  | severus_1 | + | chr3:130090881:INS:302 |  |  | Insertion | 3 | het |
|  | severus_1 | + | chr9:70898924 | - | chr9:71336169 | Deletion | 8 | het |
|  | severus_1 | + | chr9:101374815 | + | chr9:71297949 | Reciprocal inversion | 14 | het |
|  | severus_1 | - | chr9:101374847 | - | chr9:71299079 | Reciprocal inversion | 9 | het |

**S14 Impact of novel SVs on gene expression and copy number in three MPM tumour samples.** This table display transcript expression levels (TPM) versus inferred copy number variation for genes affected by novel SVs across three MPM samples (MED209, MED241, MED243). “Yes” means that gene affected by novel SV within a specific sample. The gene affected by novel SV within a specific sample represented in bold txt.

| Gene name | TPM | CN | Novel SVs | Sample |
| --- | --- | --- | --- | --- |
| BIRC6 | 14.63462 | 2 | No | MED239 |
| CDH1 | 60.16155 | 2 | No | MED239 |
| ENTR1 | 47.93122 | 2 | No | MED239 |
| EPHA7 | 0.168042 | 1 | No | MED239 |
| EXT1 | 20.71856 | 2 | No | MED239 |
| FLCN | 20.81685 | 2 | No | MED239 |
| GPC6 | 1.57654 | 1 | No | MED239 |
| JAZF1 | 11.0976 | 2 | No | MED239 |
| LPP | 7.237022 | 2 | No | MED239 |
| LRP1B | 0.008028 | 2 | No | MED239 |
| NBEA | 2.201276 | 1 | No | MED239 |
| PRKN | 0.702063 | 2 | No | MED239 |
| ROBO2 | 0.009275 | 1 | No | MED239 |
| SPECC1 | 11.22382 | 2 | No | MED239 |
| WEE1 | 10.38625 | 2 | No | MED239 |
| ANTXR1 | 129.5158 | 2 | Yes | MED239 |
| PEX5 | 8.460085 | 2 | No | MED239 |
| SH3RF3 | 3.60847 | 2 | Yes | MED239 |
| PEX5L | 16.69905 | 2 | No | MED239 |
| SULT1A2 | 18.04366 | 2 | No | MED239 |
| BIRC6 | 15.87459 | 2 | Yes | MED241 |
| CDH1 | 35.6743 | 2 | No | MED241 |
| ENTR1 | 24.30683 | 1 | Yes | MED241 |
| EPHA7 | 0.372744 | 2 | No | MED241 |
| **EXT1** | **36.74314** | **2** | **Yes** | **MED241** |
| FLCN | 15.21033 | 2 | No | MED241 |
| **GPC6** | **7.33636** | **2** | **Yes** | **MED241** |
| JAZF1 | 15.65961 | 2 | No | MED241 |
| LPP | 12.47342 | 2 | No | MED241 |
| LRP1B | 0.508839 | 2 | Yes | MED241 |
| NBEA | 2.800373 | 1 | Yes | MED241 |
| PRKN | 0.580476 | 1 | Yes | MED241 |
| ROBO2 | 0.057308 | 1 | No | MED241 |
| SPECC1 | 20.99298 | 2 | No | MED241 |
| WEE1 | 56.76337 | 2 | Yes | MED241 |
| ANTXR1 | 113.5905 | 2 | No | MED241 |
| PEX5 | 5.353839 | 2 | No | MED241 |
| SH3RF3 | 3.934655 | 2 | Yes | MED241 |
| PEX5L | 0.182746 | 2 | No | MED241 |
| SULT1A2 | 2.106599 | 2 | No | MED241 |
| BIRC6 | 12.5607 | 2 | No | MED243 |
| CDH1 | 9.654213 | 2 | Yes | MED243 |
| ENTR1 | 41.96793 | 2 | No | MED243 |
| EPHA7 | 0.111876 | 1 | Yes | MED243 |
| EXT1 | 22.66043 | 2 | No | MED243 |
| FLCN | 21.46889 | 3 | Yes | MED243 |
| GPC6 | 2.421708 | 2 | No | MED243 |
| **JAZF1** | **7.726107** | **2** | **Yes** | **MED243** |
| LPP | 5.275976 | 2 | Yes | MED243 |
| LRP1B | 0.048122 | 2 | No | MED243 |
| NBEA | 1.723013 | 1 | No | MED243 |
| PRKN | 0.607758 | 1 | No | MED243 |
| ROBO2 | 0.602227 | 2 | Yes | MED243 |
| SPECC1 | 6.613764 | 1 | Yes | MED243 |
| **WEE1** | **27.68584** | **2** | **Yes** | **MED243** |
| ANTXR1 | 107.2197 | 2 | Yes | MED243 |
| SH3RF3 | 5.201999 | 2 | Yes | MED243 |
| PEX5L | 3.287785 | 2 | Yes | MED243 |
| SULT1A2 | 7.338932 | 2 | Yes | MED243 |

| Gene name | P-value | Adjusted P-value | COSMIC | Potential cancer genes |
| --- | --- | --- | --- | --- |
| *GPC6* | **0.002746** | **0.035913** |  | **YES** |
| *PRKN* | **0.003828** | **0.043259** |  | **YES** |
| *ENTR1* | **0.000182** | **0.00687** |  | **YES** |
| *SH3RF3* | 0.030126 | 0.152034 |  | YES |
| *ANTXR1* | 0.072876 | 0.253148 |  | YES |
| *PEX5L* | 0.020906 | 0.12304 |  | YES |
| *SULT1A2* | 0.006967 | 0.063797 |  | YES |
| *WEE1* | **9.03E-06** | **0.001085** |  | **YES** |
| *LRP1B* | NA | NA | Yes |  |
| *BIRC6* | 0.050545 | 0.204197 | Yes |  |
| *EXT1* | 0.64009 | 0.822857 | Yes |  |
| *NBEA* | 0.988025 | 0.995019 | Yes |  |
| *LPP* | 0.040492 | 0.179775 | Yes |  |
| *ROBO2* | NA | NA | Yes |  |
| *EPHA7* | NA | NA | Yes |  |
| *JAZF1* | 0.586633 | 0.788334 | Yes |  |
| *CDH1* | 0.555442 | 0.768758 | Yes |  |
| *FLCN* | 0.515374 | 0.738782 | Yes |  |
| *SPECC1* | 0.017891 | 0.112493 | Yes |  |

**S15 *Table* Survival analysis of genes affected by novel SVs detected exclusively by long-read sequencing in MPM tumour samples:** the significant adjusted p-value represented in bold txt.

**S16 Table Summary of estimated immune and fibroblast cell-type fractions in three MPM tumour samples.**

|  | MED239 | | MED241 | | MED243 | | |
| --- | --- | --- | --- | --- | --- | --- | --- |
|  | Tumour | Blood | Tumour | Blood | | Tumour | Blood |
| **Fibroblasts** | 0.136 | 0.031 | 0.095 | 0.039 | | 0.352 | 0.048 |
| **Immune cells** | 0.449 | 0.929 | 0.612 | 0.926 | | 0.252 | 0.934 |
